# An Integrative Approach for Subtyping Mental Disorders Using Multimodal Data

**DOI:** 10.1101/2025.05.27.25328416

**Authors:** Yinjun Zhao, Yuanjia Wang, Ying Liu

## Abstract

Integrating multimodal biological, cognitive, and clinical data is crucial for uncovering distinct psychiatric disease subgroups, enabling precision diagnosis, personalized treatment, and more targeted drug development. However, a significant gap remains between traditional clustering approaches and the growing need for advanced methods that can integrate and jointly analyze multimodal biological and clinical data to achieve more biologically meaningful subtyping. This study introduces the Mixed INtegrative Data Subtyping (MINDS) method, a Bayesian hierarchical joint model designed to identify subtypes of Attention-Deficit/Hyperactivity Disorder (ADHD) and Obsessive-Compulsive Disorder (OCD) in adolescents using multimodal data from the Adolescent Brain Cognitive Development (ABCD) Study. MINDS integrates clinical assessments, neuro-cognitive measures, and neuroimaging biomarkers while simultaneously performing clustering and dimension reduction. By leveraging Polya-Gamma augmentation, we propose an efficient Gibbs sampler to improve computational efficiency and provide subtype identification. Simulation studies demonstrate the superior robustness of MINDS compared to traditional clustering techniques. Application to the ABCD study reveals more reliable and clinically meaningful subtypes of ADHD and OCD with distinct cognitive and behavioral profiles. These findings show the potential of multimodal model-based clustering for advancing precision psychiatry in mental health.

## 1. INTRODUCTION

Over the past four decades, considerable efforts have been devoted to applying unsupervised machine learning algorithms to dissect the heterogeneity of mental disorders and discover subtypes (Schnack, 2019; Marquand et al., 2016). Unfortunately, the lack of reproducibility in clustering algorithms has been a major obstacle for these methods to provide clinically useful subtyping for any mental disorder (Marquand et al., 2019). Traditional methods for mental disorder subtyping often rely on a single data type, such as clinical symptoms or neuroimaging measures, which may fail to accurately capture the complex, overlapping functional domains of mental health conditions or their underlying biological and psychological mechanisms (Kozak and Cuthbert, 2016).

The Research Domain Criteria (RDoC) initiative, launched by the National Institute of Mental Health, proposes to integrate biological and behavioral measures from various sources of analysis and different domains of functioning to create dimensional latent constructs of mental disorders and improve the precision of diagnosis and treatment (Insel et al., 2010). The Adolescent Brain Cognitive Development (ABCD) study is the largest and most comprehensive nationwide initiative in the United States dedicated to research on adolescent brain and behavioral development. By collecting multimodal data, including neuroimaging, behavioral tasks, clinical evaluations, and lifestyle factors, the ABCD study captures a comprehensive view of the biological and environmental influences during the critical developmental years of adolescence. This rich study presents an unprecedented opportunity to address the heterogeneity of mental health conditions through unsupervised learning techniques, such as clustering. In this paper, we fill a gap in mental health research by introducing a novel method that integrates multimodal data of different distribution types while simultaneously performing clustering and dimension reduction. Our goal is to develop integrative subtyping techniques to identify Attention-Deficit/Hyperactivity Disorder (ADHD) and Obsessive-Compulsive Disorder (OCD) subtypes in adolescents in the ABCD study. To achieve this, we follow the RDoC paradigm to integrate individual items from clinical assessments, such as the Kiddie Schedule for Affective Disorders and Schizophrenia (KSADS) modality, and neuro-cognitive measures from the NIH Cognitive Toolbox, along with cognitive task data and cortical thickness in selected brain regions.

Conventional clustering and subtyping methods have several limitations in integrating multimodal data and identifying meaningful subtypes. For example, distance-based approaches such as K-means and hierarchical clustering are widely applied in mental health research. However, they rely on predefined distance measures that may be sensitive to noise and outliers to form clusters, which reduces their statistical robustness (Chen et al., 2022). Additionally, they lack the capability to estimate latent constructs, which is essential in RDoC to reveal patterns of measures of mental disorders and capture core mental functions (Lanza et al., 2007). Model-based subtyping methods, such as latent class analysis (Magidson and Vermunt, 2002), normative models (Marquand et al., 2016), and finite mixture models (McLachlan, 2000), are generally considered more robust than distance-based clustering because they use probabilistic frameworks to account for uncertainty and the distribution of data (Little, 2013), rather than arbitrary distance measures. By modeling the data within probabilistic frameworks, one can enhance the reliability and reproducibility of identifying clinically meaningful subtypes. However, these models are typically designed to handle a single type of data distribution and cannot integrate multiple modalities or data types (e.g., neuroimaging, clinical symptoms, and behavioral data), limiting their applicability in studies that require a comprehensive, multi-modal analysis (Sinha et al., 2021).

To accommodate mixed-type data, distance-based subtyping methods can be adapted to use different similarity measures for each type (e.g., Euclidean for continuous variables, Dice or Jaccard for binary variables). For instance, Gower’s distance computes similarity across mixed data types and is widely used for this purpose (Gower, 1971). Another common approach for clustering mixed-type data is a two-step feature-based method. The first step involves feature transformation with or without dimension reduction, such as binary-to-continuous transformation (e.g., using logistic regression to convert binary variables into probability to estimate liability scores (McLachlan, 2000)), factor analysis with polychoric or polyserial correlations, or principal component analysis (PCA) with Gower’s distance, or Joint and Individual Variation Explained (JIVE) with low-rank variance decomposition for integrated data (Lock et al., 2013). This creates a compact, continuous representation of the mixed-type data. The second step applies conventional clustering to these transformed features. iClusterBayes also adopts a two-step approach to integrating and clustering mixed data types. First, it jointly models continuous and binary variables to project them into a dimension-reduced latent space. Then, individuals are clustered within this latent space using the K-means algorithm (Mo et al., 2018). While these methods leverage the strengths of each data type, using only intermediate features instead of the original data for clustering may lead to the loss of shared information and reduce efficiency. Latent class analysis (LCA) can jointly model both continuous and categorical/binary modalities; however, fitting LCA models may pose computational challenges because the joint likelihood (e.g., for combining multinomial and Gaussian distributions) often lacks a closed-form solution. Numeric integration is required in the estimation process, which becomes infeasible when the dimension of the latent constructs is high (Hagenaars and McCutcheon, 2002).

In this paper, we develop a Mixed INtegrative Data Subtyping (MINDS) method for subtyping mental disorders and clustering individuals by integrating measures across mixed-type modalities. We propose Bayesian hierarchical joint models with latent variables based on item response theory (IRT) and finite mixture modeling. For models integrating categorical data such as KSADS, we utilize Pólya-Gamma augmentation (Polson et al., 2013), which converts the logistic likelihood into a conditionally conjugate form. This transformation enables efficient Gibbs sampling and makes it feasible and efficient to handle the integration of diverse data types within a Bayesian framework. Our method tackles several challenges in clustering multimodal data. First, MINDS can handle diverse data types, such as categorical and continuous, into a unified clustering model. Second, beyond assigning cluster membership, MINDS estimates latent constructs for each individual to assist in explaining the unique characteristics of each cluster. This is essential for disease subtyping, identifying subject-specific disease risks, and developing targeted interventions. Third, our model extends the IRT to perform clustering and dimension reduction at the same time, which improves both interpretability and computational efficiency. Fourth, we propose to use Pólya-Gamma augmentation to facilitate Gibbs sampling, which is computationally more efficient than many other Bayesian methods. Finally, MINDS provides a flexible framework for handling missing data, allowing all observed measures from a subject to be included in the likelihood for estimation. Some other approaches may require complete data across all variables or rely on imputation prior to the analysis.

The rest of this paper is organized as follows. Section 2 introduces the proposed models and Pólya-Gamma augmentation for integrating categorical/binary modality, as well as the details of the algorithm. Section 3 presents simulation studies conducted to assess the consistency of the estimator and the efficiency of our method. Here, we also compare the performance of our approach to several alternative methods. In Section 4, we apply our methods to the ABCD study, subtyping adolescent participants based on symptoms, neuro-cognitive measures, and brain measures related to ADHD and OCD. We conclude with limitations and extensions in Section 5.

## 2. MODEL

### 2.1. The IRT model

The Item Response Theory (De Ayala, 2013) is a framework used to model the relationship between latent traits, i.e., unobserved characteristics or attributes, such as attention or cognitive control, and their manifestations in observed variables, such as responses to items in questionnaires or clinical instruments. The IRT is widely used in educational testing, psychology, psychometrics, and other fields where assessments and questionnaires are utilized to measure underlying traits, including ability, attitude, or personality (Van Der Linden and Hambleton, 1997). It is a suitable model to capture the relationship between binary responses in the KSADS to identify latent traits of attention and cognitive control. MINDS addresses multimodal integration by extending the IRT with Bayesian hierarchical joint modeling for clustering and integrating different data types. In the following, we briefly introduce commonly used IRT models and how they are adapted in MINDS.

In the Rasch model, for a binary item, the probability that a person *i* with a latent trait *θ*_*i*_ (e.g., attention) endorsing item *j* with difficulty parameter *δ*_*j*_ is given by a logistic regression with random effects,

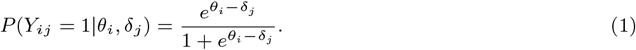

The relationship specified in Equation (1) is also called the item response function (IRF). To accommodate the non-unity slope of the IRF, an additional parameter *α* is introduced to generalize the Rasch model to a one-parameter logistic (1PL) model such that

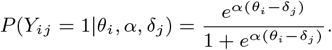

When *α* varies across items, the 1PL model becomes a two-parameter logistic (2PL) model as

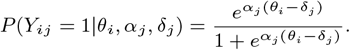

An item’s *α*_*j*_ characterizes how well the item can differentiate among individuals located at different points on the continuum. Thus, *α*_*j*_ is also referred to as the item *j*’s discrimination parameter.

The multidimensional IRT (MIRT) model extends the traditional unidimensional IRT to account for multiple latent traits, such as attention and cognitive control, as

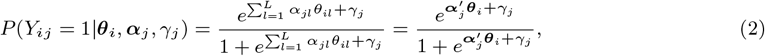

where ***θ***_*i*_ = {*θ*_*il*_}_*l*=1,…,*L*_ is the *L*-dimensional vector of latent traits for person *i*, ***α***_*j*_ = {*α*_*jl*_}_*l*=1,…,*L*_ is the *L*-dimensional vector of discrimination parameters for item *j*, and 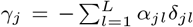 is the difficulty parameter for item *j*, also referred to as the item *j*’s threshold. This is useful when test items are conceptualized to assess multiple underlying abilities or attributes.

### 2.2. Latent mixture model with single modality

The conventional MIRT models can only conduct dimension reduction for categorical data and do not accommodate mixture distributions. Here, we extend them to incorporate continuous variables and conduct clustering simultaneously. We first describe the modeling for binary, ordinal, or continuous measures, respectively, and then introduce how to integrate different data types.

Let *Y*_*ij*_ denote the binary modality’s *j*th item in a clinical instrument from the *i*th subject, where *i* = 1, …, *N*_*b*_, and *j* = 1, …, *N*_*d*_. We propose a latent mixture model inspired by MIRT for the binary modality as

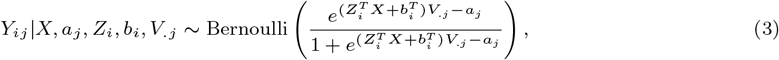

where *X* is a *N*_*c*_ × *N*_*t*_ matrix representing the *N*_*c*_ cluster centers, each of which is a *N*_*t*_-dimensional vector. Here, *Z*_*i*_ is an indicator of the *i*th subject’s unobserved cluster membership assumed to follow a categorical distribution, *p*(*Z*_*i*_ = *e*_*k*_) = *θ*_*k*_, where *e*_*k*_ is the *k*-th standard basis vector in ℝ^*K*^, *i*.*e*., *e*_*k*_ = (0, 0, …, 1, …, 0)^⊤^ with the 1 in the *k*-th position. *b*_*i*_ is the *i*th subject’s latent construct, e.g., cognitive control ability that deviates from his or her cluster mean ability. Furthermore, *V*_.*j*_ is an *N*_*t*_-dimensional loading vector on latent construct for the *j*th item representing how well the items can differentiate among individuals at different points of latent constructs, and *a*_*j*_ represents the *j*th item’s threshold. Note that the above model reduces to the MIRT model when there is a single cluster. A similar method under a cumulative model can be developed for ordinal measures as

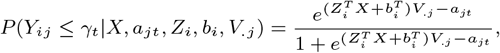

where 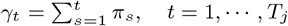, …, *T*_*j*_, and *π*_*t*_ represents the probability that *Y*_*ij*_ belongs to *t*th category, and *a*_*jt*_ is the threshold parameter for *j*th item belonging to *t*th category. For modeling *N*_*d*_-dimensional continuous measures *Y*_*ij*_, we propose a multivariate Gaussian model as

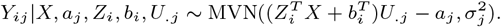

The interpretations of *a*_*j*_, *Z*_*i*_, *b*_*i*_, *U*_.*j*_ are similar to *X, a*_*j*_, *Z*_*i*_, *b*_*i*_, *V*_.*j*_ in model (3). In these models, individuals in different subtypes have distinct mean latent traits (i.e., cluster center *X*) while sharing other parameters such as discriminant and threshold values.

Since the binary modality’s distribution does not belong to the Gaussian family, the posterior distribution is not conjugate to the prior, making Gibbs sampling inapplicable. Pólya-Gamma augmentation has been proposed to yield a Gibbs sampler for the Bayesian logistic model (Polson et al., 2013). A random variable *Y* has a Pólya-Gamma distribution with parameters *b >* 0 and *c* ∈ ℝ, denoted as *Y* ∼ PG(*b, c*), if

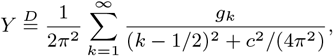

where *g*_*k*_ ∼ Ga(*b*, 1) are independent gamma random variables, and 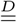 indicates equality in distribution. Let *p*(*ω*) denote the density of the random variable *ω* ∼ PG(*b*, 0), *b >* 0. Then the following integral identity holds for all *a* ∈ ℝ:

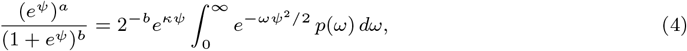

where *κ* = *a* − *b/*2. Moreover, the conditional distribution *p*(*ω*|*ψ*), where *ψ* is the exponential part in (4), is also in the Pólya-Gamma class, i.e.,

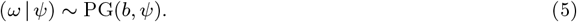

The properties of (4) (5) suggest a simple strategy for Gibbs sampling across a wide class of binomial models, that is, Gaussian draws for the main parameters and Pólya-Gamma draws for a single layer of latent variables. Under model (3), the conditional distribution of *Y* given all the rest of parameters is

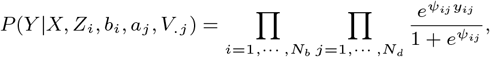

By Equation (4), the conditional distribution can be expressed as

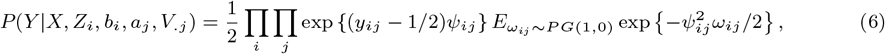

where 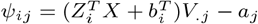.

Given *ω*_*ij*_, the expectation in Equation (10) is exp 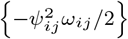, which leads the conditional probability of *Y*_*ij*_ in model (3) follows the Gaussian distribution.

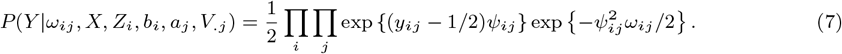

By the property of the Pólya-gamma distribution in Equation (5), we have

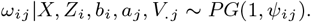

In this sense, all parameters’ posterior distributions are conjugate to their prior distributions, so the Gibbs sampling can be adopted for parameter estimation.

### 2.3. Latent mixture model with multi-modalities

Model (3) only handles a single modality. Here, we extend the single-modality model to handle more complex multimodal data using discrete clinical and continuous behavioral measures. We propose a joint model that integrates binary and continuous modalities as follows

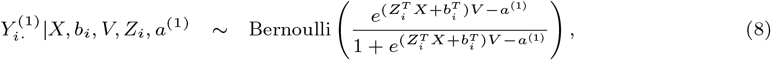

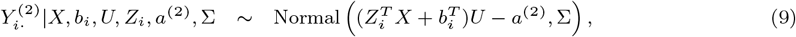

where *X* is a matrix of cluster centers shared by these two modalities, *Z*_*i*_ is subject *i*th’s cluster membership, *b*_*i*_ is *N*_*t*_-dimensional subject’s latent construct, *V* and *U* are 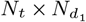 -dimensional and 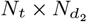-dimensional loading matrix on cluster center *X* for binary and continuous modality, respectively. 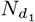 represents the number of items in binary modality, and 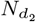 represents the number of measures in continuous modality. *a* ^(1)^ and *a* ^(2)^ represent item difficulty for two different modalities. For continuous modality, Σ is a diagonal covariance matrix with diagonal entries 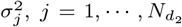. In this model, we assume the two data-type modalities share the cluster center *X*, and each subject belongs to the same cluster under different modalities.

We use a Bayesian hierarchical algorithm to estimate the parameters. We assume the following prior distributions for the parameters in (8) and (9) as that *Z*_*i*_ follows categorical distribution with hyperparameter *θ*; *b*_*i*_ follows 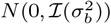; *X, V, U* are from multivariate normal distributions, 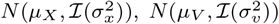, and 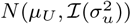, respectively; *a*^(1)^, *a*^(2)^ are from multivariate normal, 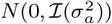, respectively. We assume *θ* follows Dirichlet distribution with parameters of equal weight 1*/N*_*c*_, and 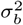 and 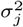 follow Inverse Gamma distribution *IG*(*α*_*b*_, *β*_*b*_) and *IG*(*α*_*j*_, *β*_*j*_), respectively.

The conditional distribution of *Y* given all the rest parameters in the joint model (8)-(9) is

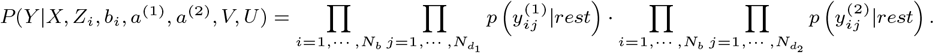

A similar method is used as for model (3), which introduces a latent variable *ω* following the Pólya-Gamma distribution such that 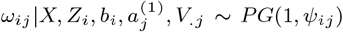, where 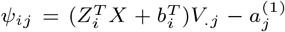. Similar to equation (11), the conditional distribution of 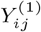 given *ω* and all the rest of the parameters in model (8) is from a Gaussian family. In this sense, the posterior distributions of all the parameters 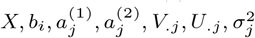 in the joint model are conjugate to their prior distributions. The conditional probability that the *i*th subject belongs to the *k*th cluster, given the rest of the parameters, denoted as *π*_*ik*_, is expressed as:

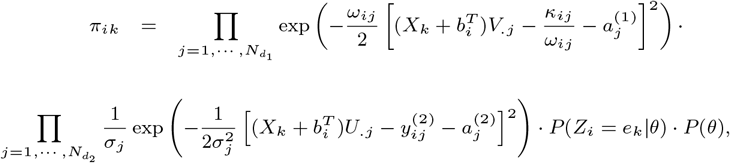

which indicates that the posterior distribution of *Z*_*i*_ is from Multi(1, *π*_*i*_), 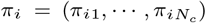. All the parameters’ posterior distributions can be expressed in closed forms so that the Gibbs sampling can be adopted for parameter estimation. Details of the posterior distributions are in section 2.4.

#### Algorithm 1

Gibbs sampling steps for joint model

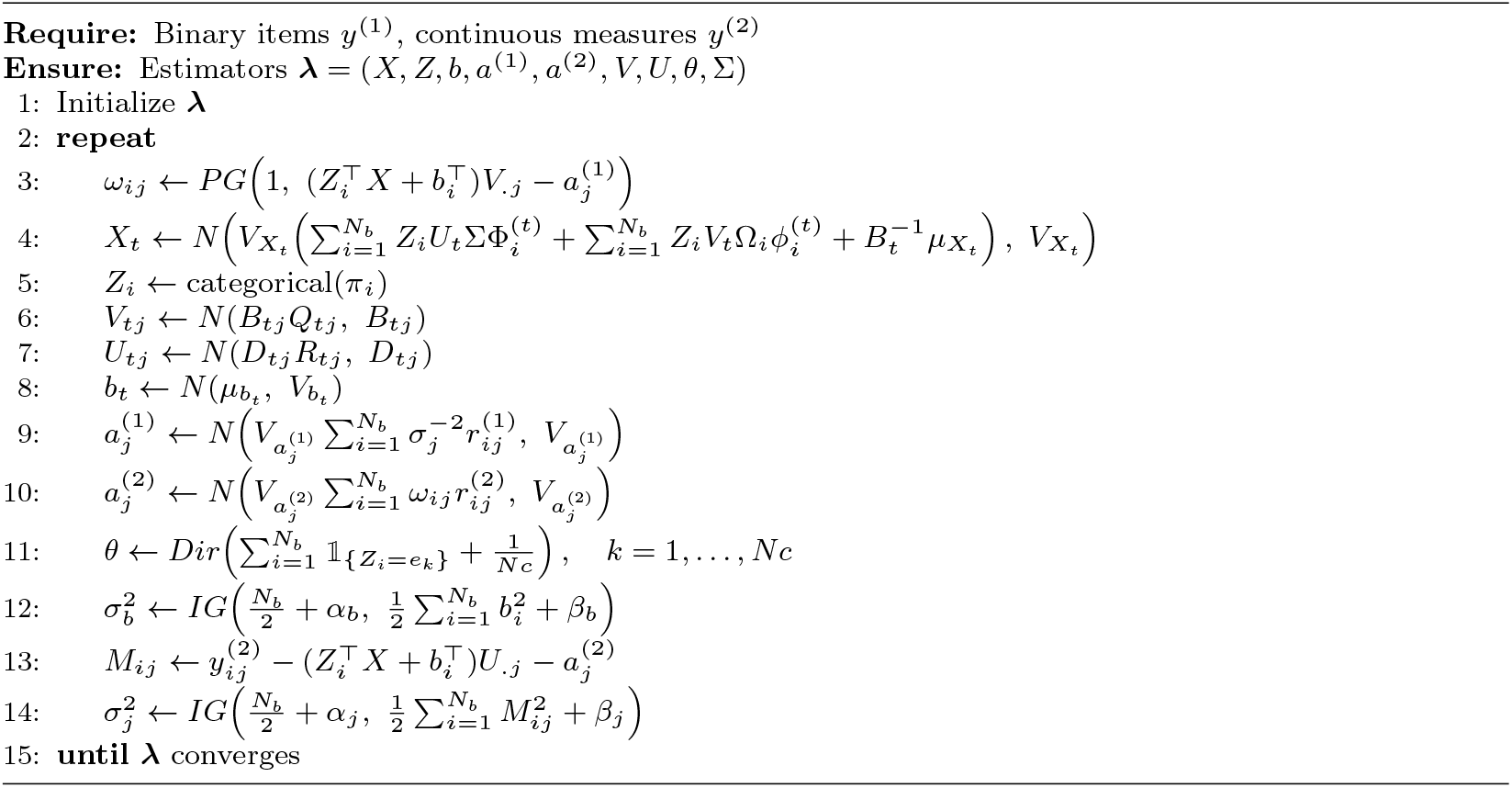

### 2.4. Algorithm

We derive Gibbs sampling steps for the joint model (8)&(9) in Algorithm 1. This algorithm can be easily modified to accommodate the single modality model (3), a simpler version of the joint model. Information Criterion (IC), a selection information criterion for Bayesian models (Ando, 2011), which avoids the problems of over-fitting associated with DIC (Spiegelhalter et al., 2014), is used to determine the optimal number of clusters *N*_*c*_.

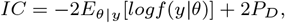

where *P*_*D*_ is defined as difference between the posterior mean of deviance and the deviance estimated at the posterior mean of the parameters, 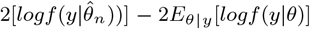, and 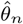 is the posterior mean. The stationary trace plot is used to examine convergence.

In Algorithm 1, for step 4,

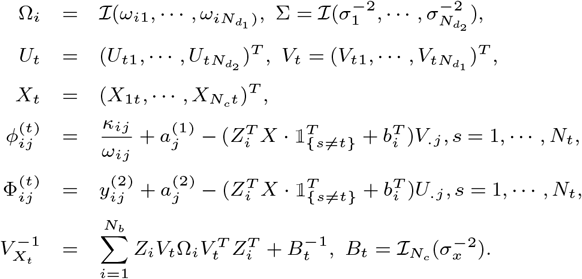

For step 5, the conditional probability of *i*th subject belongs to the *k*th cluster center, i.e., 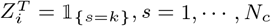, *s* = 1, …, *N*_*c*_, given the rest parameters is

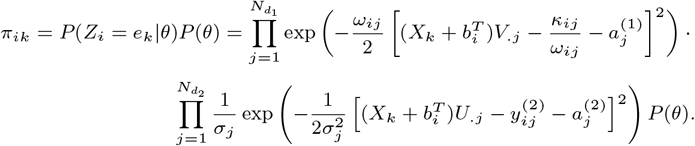

For steps 6 and 7,

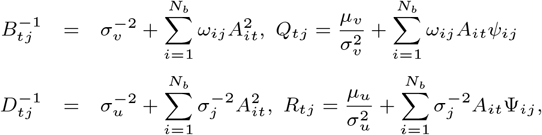

where

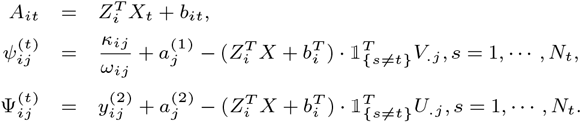

For step 8,

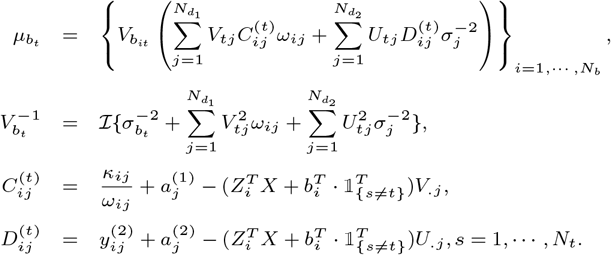

For steps 9 and 10,

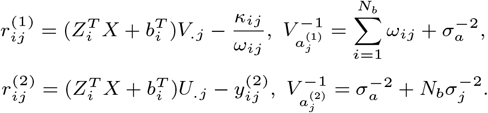

This model has metric indeterminacy in the sense that we do not have a metric for *V* and *U* with an intrinsic origin or unit. In addition, the model has a rotational indeterminacy in the sense that the axes of *X* that represent latent variables are free to rotate about their origin because there is no external reference to fix their orientation. To address these issues and ensure identifiability, we set *X*_11_ to be one and 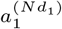 to be zero and estimate *X, U* and *V* by solving equation system that

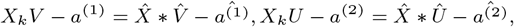

where 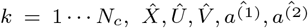 are estimation from algorithm 1. The equation system has unique solution when the number of parameters, 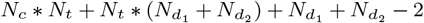 is not greater than the number of equations, 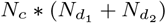, i.e. 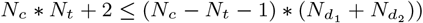.

## 3. SIMULATION STUDIES

We conducted extensive simulation studies to evaluate the performance of our proposed algorithm for the joint model. This involved examining the consistency of latent variable estimates, calculating training error on the training data and test errors on testing data when the cluster membership is known, and comparisons with alternative methods.

We generated three simulation settings, each setting composed of 10 continuous measures and 10, 20, 30 binary items, respectively. Three latent constructs were considered for the cluster center *X* and the *i*th subject’s latent construct *b*_*i*_, i.e., *N*_*t*_ = 3. The elements of the true cluster center *X* and subject-specific ability *b*_*i*_ were randomly drawn from a uniform distribution *U* (0, 2) and a Gaussian distribution *N* (0, 0.2), respectively. Subjects were generated from five clusters, i.e., *N*_*c*_ = 5, with cluster membership weights *θ* to be (0.3, 0.15, 0.15, 0.2, 0.2), based on which we generated true membership *Z*_*i*_ from Categorical(*θ*) for subject *i*. The item difficulty parameters 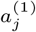 for binary items and 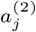 for continuous measures were randomly drawn from U(-0.5, 0.5) and U(-5, 5), respectively. All elements of the loading matrix *V* for binary items and *U* for continuous measures were independently randomly drawn from U(0, 2). All the prior distributions were non-informative, and all the estimators’ initial values were random.

We conducted 200 replications for each setting and sample size (1,000 and 2,000) to evaluate the consistency of latent variable estimates and training error. As shown in Web Figures 1-4, the medians of the estimated values for cluster center *X*, loading matrix *U* and *V*, and item difficulty *a*^(1)^ and *a*^(2)^ are close to their true values except for *U*_13_, *V*_15_. As the sample size increases from 1000 to 2000, the variance of the estimators decreases. We compared the root mean squared errors (RMSE) and bias of all parameters for sample size 1,000 and 2,000. As shown in Table 1, overall parameters’ RMSEs decrease as the sample size increases from 1,000 to 2,000. The other two settings have similar performance, and results are omitted.

**Table 1.** Simulation results from a joint model based on 200 replicates; RMSE and bias are calculated for 1,000 vs. 2,000 training sample size; each model integrated one binary and one continuous modality, incorporating 10–30 binary items and 10 continuous measures.

| | Training Size | Items | $x$ | $V$ | $U$ | $a$ | $\theta$ |
| --- | --- | --- | --- | --- | --- | --- | --- |
| BIAS | 1000 | 10 | 0.054 | 0.046 | 0.024 | 0.273 | 0.006 |
|  | 2000 | 10 | 0.059 | 0.036 | 0.024 | 0.271 | 0.003 |
|  | 1000 | 20 | 0.093 | 0.055 | 0.032 | 0.252 | 0.012 |
|  | 2000 | 20 | 0.084 | 0.042 | 0.026 | 0.244 | 0.009 |
|  | 1000 | 30 | 0.076 | 0.059 | 0.062 | 0.194 | 0.01 |
|  | 2000 | 30 | 0.056 | 0.042 | 0.034 | 0.187 | 0.001 |
| RMSE | 1000 | 10 | 0.158 | 0.156 | 0.09 | 0.314 | 0.008 |
|  | 2000 | 10 | 0.134 | 0.117 | 0.074 | 0.303 | 0.005 |
|  | 1000 | 20 | 2.087 | 0.647 | 0.691 | 1.457 | 0.836 |
|  | 2000 | 20 | 2.092 | 0.651 | 0.688 | 1.458 | 0.836 |
|  | 1000 | 30 | 2.17 | 0.808 | 0.578 | 1.239 | 0.853 |
|  | 2000 | 30 | 2.161 | 0.808 | 0.571 | 1.243 | 0.863 |

**Fig. 1.**
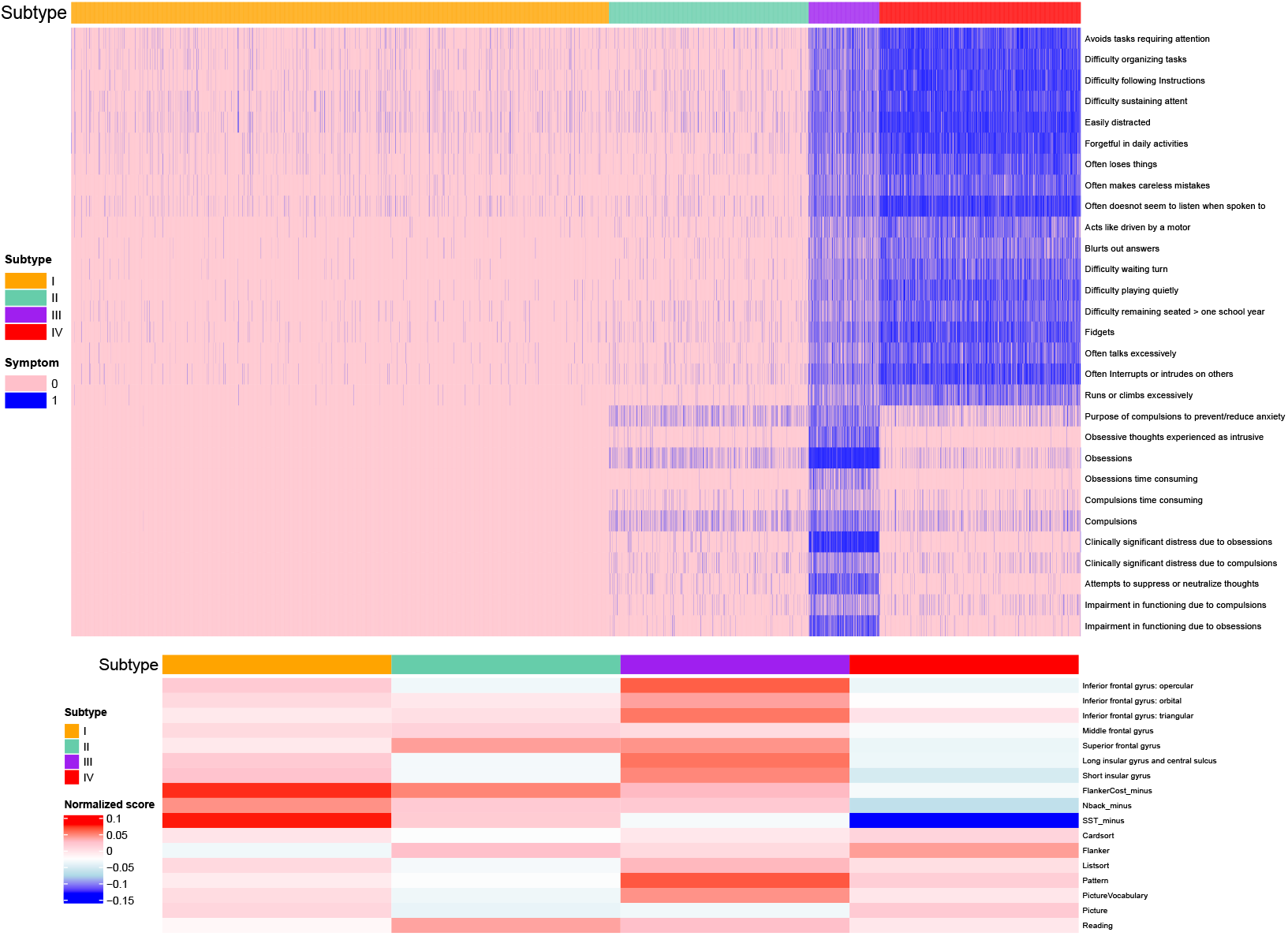
Heatmaps of binary items and continuous measures across subtypes by the MINDS method, integrating ADHD- and OCD-related symptoms with selected neuro-cognitive measures. The top heatmap includes eighteen binary KSADS ADHD symptoms and eleven binary KSADS OCD symptoms. Bottom heatmap includes seven cortical thickness measures in selected brain regions, seven NIH Toolbox cognition assessments, and three behavioral indices specifically targeting attention and inhibitory control; scores are normalized by row.

Corresponding to the sample size range of the motivating ABCD study, 10,000 subjects were generated as the test data to assess the testing error by using the same true values of parameters *X, U, V, a*^(1)^, and *a*^(2)^ as those used in the generation of the training data. For the subject-specific parameters *Z*_*i*_ and *b*_*i*_, the same distributions with the same parameters *θ* and 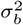 as in the training data were used in the generation of the testing data, respectively. To get the predicted cluster membership, we plugged the estimated values of *X, U, V, a*^(1)^, and *a*^(2)^ from training data, and iterated the remaining unknown parameters *ω, Z*, and *b*_*i*_ until convergence. The estimated *Z* returns the predicted cluster membership. The test errors were calculated using three metrics to evaluate the performance of the clustering. The three metrics are (a) Bayes error rate, which is defined as the expectation of the event that the predicted category (the one having maximum membership weight) is not equal to the true category; (b) Classification error, the proportion that the predicted category is not equal to the true category; (c) Jaccard distance, defined as the one minus the size of the intersection divided by the size of the union of two label sets. For all three metrics, smaller scores imply better performance.

We compared the results of the MINDS method with six other alternative approaches: iClusterBayes, K-means clustering, Hierarchical clustering (shortened for Hclust), two-step JIVE, two-step K-means clustering, and two-step Hclust. In two-step JIVE, principal component (PC) scores were obtained by JIVE, and then K-means was used to cluster the PC scores. In the two-step K-means, we first conducted factor analysis based on the polychoric correlation, and in the second step, used K-means to cluster the factor scores into *N*_*c*_ clusters. The two-step Hclust differs from the two-step K-means in that, in the second step, it uses Hclust to cluster the factor scores. As shown in Table 2 3 and Web Table 1, the training and testing errors of MINDS are lower than all the other six methods in all three metrics, which shows that the performance of MINDS is satisfactory.

**Table 2.** Mean(sd) of training set Bayes error from MINDS and alternative methods, evaluated across 200 replicates. Training sample sizes ranged from 1,000 to 2,000 subjects. Each model integrated one binary and one continuous modality, incorporating 10–30 binary items and 10 continuous measures.

| Items | Training Size | Training Set Bayes Error |  |  |  |  |  |  |
| --- | --- | --- | --- | --- | --- | --- | --- | --- |
|  |  | MINDS | iClusterBayes | Two-step JIVE | K-means | Hclust | Two-step K-means | Two-step Hclust |
| 10 | 1000 | 0.049(0.03) | 0.107(0.01) | 0.135(0.009) | 0.111(0.044) | 0.152(0.01) | 0.107(0.042) | 0.134(0.032) |
| 10 | 2000 | 0.046(0.024) | 0.104(0.008) | 0.141(0.006) | 0.127(0.038) | 0.15(0.009) | 0.12(0.043) | 0.145(0.031) |
| 20 | 1000 | 0.028(0.021) | 0.081(0.015) | 0.168(0.026) | 0.084(0.062) | 0.144(0.034) | 0.089(0.036) | 0.107(0.026) |
| 20 | 2000 | 0.028(0.021) | 0.072(0.013) | 0.169(0.032) | 0.085(0.064) | 0.141(0.027) | 0.084(0.038) | 0.101(0.028) |
| 30 | 1000 | 0.033(0.013) | 0.086(0.017) | 0.198(0.006) | 0.087(0.056) | 0.108(0.01) | 0.097(0.039) | 0.105(0.031) |
| 30 | 2000 | 0.035(0.016) | 0.087(0.016) | 0.206(0.004) | 0.099(0.054) | 0.109(0.012) | 0.093(0.037) | 0.129(0.041) |

**Table 3.**
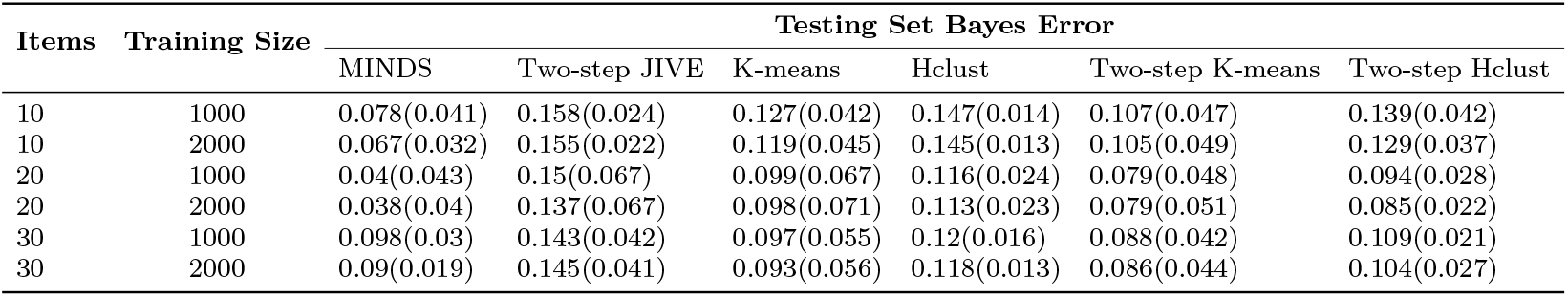
Mean(sd) of testing set Bayes error from MINDS and alternative methods, evaluated across 200 replicates. Training sample sizes ranged from 1,000 to 2,000 subjects. The testing sample includes 10,000 subjects. Each model integrated one binary and one continuous modality, incorporating 10–30 binary items and 10 continuous measures.

The computing time for one iteration of a joint model with 2,000 samples when the number of binary items varies from 100, 200, and 400 items was 0.7, 1.2 and 2.2 seconds, respectively, by Apple M1 Pro.

## 4. DISEASE SUBTYPING IN THE ABCD STUDY

We applied MINDS to identify subtypes of subjects in the ABCD study by jointly integrating ADHD- and OCD-related symptoms with neurocognitive behavioral and brain measures. The analysis incorporated multiple modalities: eighteen binary KSADS ADHD symptoms, eleven binary KSADS OCD symptoms, seven continuous brain measures, and seven NIH Toolbox cognition assessments. The seven brain measures were the mean cortical thickness values averaged across the right and left hemisphere values. The regions of interest (ROIs) are defined by the Destriuex atlas Destrieux et al. (2010). The T1-weighted images were preprocessed using the FreeSurfer 5.1 pipeline Fischl et al. (1999). We consider 7 ROIs which are found relevant to ADHD and OCD diagnosis: the opercular, orbital, and triangular parts of the inferior frontal gyrus; the middle and superior frontal gyri; the long insular gyrus; and the central sulcus of the insula(Norman et al., 2017; Brem et al., 2014; Thorsen et al., 2020). For each region. Among baseline participants aged 9–11 years, greater cortical thickness is considered favorable and reflects healthier neuro-development. The Toolbox tasks included Picture Vocabulary, Flanker, List Sort, Card Sort, Picture Sequence, Reading, and Pattern Comparison, which capture a range of cognitive domains such as language, reading, working memory, attention, and response inhibition (Thompson et al., 2019). The higher the Toolbox cognitive scores, the better the performance in neuro-cognitive development. In addition, we included three behavioral indices specifically targeting attention and inhibitory control (Casey et al., 2018; Siemann et al., 2018). These were: (i) the Flanker Cost Effect (mean reaction time for incongruent trials minus that for congruent trials), indexing interference suppression; (ii) the N-back Score (standard deviation of reaction time divided by mean reaction time in the EN-back task), indexing working memory stability; and (iii) the Stop Signal Task (SST) Score (standard deviation of reaction time divided by mean reaction time in the SST), indexing inhibitory control consistency. A detailed description of all cognitive measures is provided in Table 4.

**Table 4.** Neuro-cognitive measures used in the ABCD study participants subtyping.

| Variable | Task/Test | What it measures |
| --- | --- | --- |
| SST Score | The Stop Signal Task | Response inhibition |
| Nback Score | The EN-back Task | Working memory |
| Flanker Cost Effect | Toolbox Flanker Task | Attention, inhibition |
| Pic Vocab | Toolbox Picture Vocabulary Task | Language |
| Flanker | Toolbox Flanker Task | Attention, inhibition of automatic response |
| List Sort | Toolbox List Sorting Working Memory Test | Working memory, information processing |
| Card Sort | Toolbox Dimensional Change Card Sort Task | Executive function |
| Pattern | Toolbox Pattern Comparison Processing Speed Test | Information processing |
| Picture | Toolbox Picture Sequence Memory Test | Episodic memory, sequencing |
| Reading | Toolbox Oral Reading Recognition Task | Language, reading/decoding skills |

We analyzed participants at baseline, excluding 152 individuals with missing DSM ADHD or OCD diagnoses and 548 individuals with complete missingness in any of the four modalities out of 11,878 total participants, as those with complete missingness in a modality might have systematically different distributions. This yielded a final analytic sample of 10,126 participants. In model fitting, each of the three variability scores was negated so that higher values reflected greater child ability, consistent with the direction of the Toolbox and brain measures. The four-group solution was selected based on the lowest information criterion. Four distinct subtypes were identified by integrating ADHD- and OCD-related symptoms with cortical and cognitive profiles, as illustrated in Figure 1. Subtype I (Healthy) represented individuals with mild ADHD inattention symptoms and minimal ADHD impulsivity and hyperactivity symptoms. They showed only slightly reduced cortical thickness and relatively intact cognition, with mild deficits in reading and attention, accompanied by efficient attentional control, working memory, and stable inhibition. Subtype II (Mild Symptomatic with Cognitive Difficulties) displayed mild ADHD and OCD symptoms, but substantial cortical thinning across multiple regions. This group demonstrated broad cognitive difficulties across domains, except for Reading and Flanker, while their attentional control and inhibition remained moderately efficient. Subtype III (ADHD&OCD-dominant) was characterized by severe ADHD symptoms (inattention, impulsivity, and hyperactivity) across all items, alongside pervasive OCD symptoms (obsession and compulsion) and inefficiency in stable inhibition as indexed by SST tasks. Despite this, these individuals exhibited preserved cortical thickness and relatively strong cognitive performance across most NIH Toolbox domains. Finally, Subtype IV (ADHD-dominant with Reduced Brain Development) was marked by severe ADHD symptoms, mild OCD features, and pronounced cortical thinning. Unlike Subtype III, this group showed above-average performance in most cognitive domains, yet exhibited reduced stability of attentional control, working memory, and inhibition.

To compare MINDS with DSM diagnosis, which is the standard criterion for diagnosing ADHD and OCD, we calculated the Calinski-Harabasz Index (CH) as

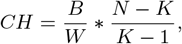

where *B* is the sum of squares between clusters, *W* is the sum of squares within clusters, *N* is the total number of data points, *K* is the number of clusters. A higher score of CH indicates larger between-cluster variation and smaller within-cluster variation. The MINDS method is better at capturing homogeneity and distinguishing heterogeneity between clusters than clustering by ADHD/OCD DSM diagnosis, which groups the participants into ADHD&OCD, ADHD, OCD, and None. The Calinski–Harabasz indices (CHs) for continuous measures were 3.45 for the MINDS method versus 1.73 for the ADHD/OCD DSM diagnosis, indicating that MINDS performed 99% better. For symptom data, the CHs were 3490 for MINDS compared with 1308 for DSM, representing a 166% improvement.

To examine the external validity of the subtypes from MINDS, Figure 2 shows the distribution of self-reported grades by parents among these subtypes in the third follow-up year. Our analysis indicates that Subtype I (Healthy) and II (Mild Symptomatic with Cognitive difficulties) exhibit the highest academic achievement. In contrast, Subtype III (ADHD&OCD-dominant) and IV (ADHD-dominant with Reduced Brain Development) demonstrate the lowest academic performance. These results were further confirmed by the results (as shown in Figure 3) of the proportional odds logistic regression of the third-year self-reported grades on participants’ subtypes adjusted by age, gender, race, education, family income, etc. Subgroup III (OR: 1.45, 95% CI: 1.14-1.84) and Subgroup IV (OR: 2.54, 95% CI: 2.16-2.98) are both significantly associated with poor academic performance compared to the Healthy group.

**Fig. 2.**
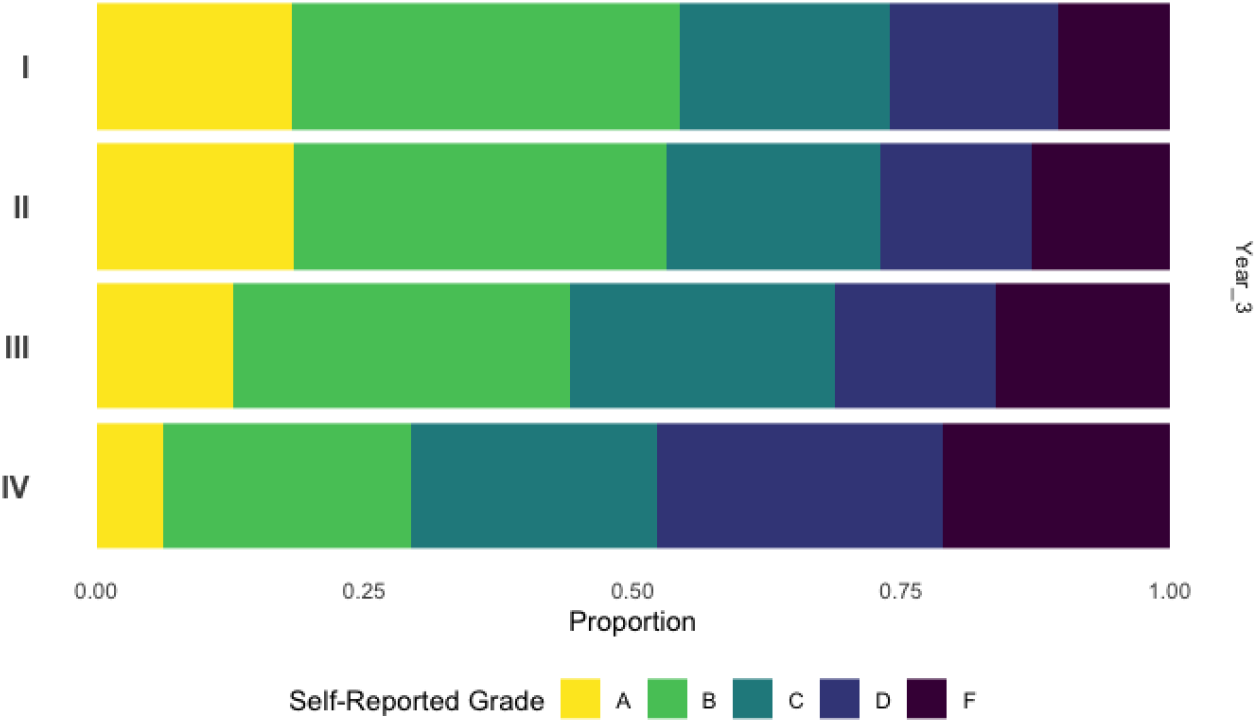
Proportion of grades self-reported by parents in different subtypes at the third follow-up year.

**Fig. 3.**
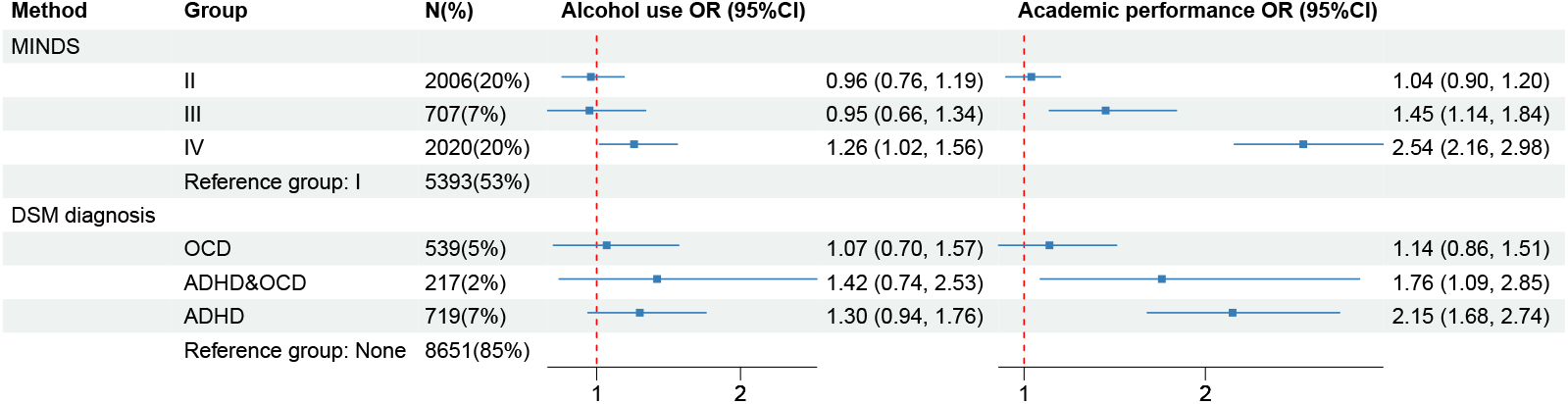
Associations of subtypes with alcohol use and poor academic performance, expressed as odds ratios (95% CI) versus the healthy group, based on the MINDS method and DSM ADHD&OCD diagnoses.

Interestingly, Subtype II, despite exhibiting mild OCD symptoms and broad cognitive difficulties across most Toolbox domains, demonstrated relatively strong academic performance. Notably, it achieved the highest reading score among all subtypes, which may serve as a compensatory factor supporting its overall academic achievement.

The DSM diagnosis subtyping also finds two significant groups, the ADHD&OCD group (OR: 1.76, 95%CI: 1.09-2.85), and the ADHD group(OR: 2.15, 95%CI: 1.68-2.74) associated with academic performance as shown in Figure 3. However, MINDS identified 27% of subjects, compared with only 9% by DSM diagnosis, as being at risk for poor academic performance and in need of psychological and educational interventions.

Additionally, MINDS results reveal that the Subtype IV has significantly higher odds of alcohol use compared to the Healthy group (OR: 1.26, 95%CI: 1.02-1.56), shown in Figure 3, whereas the DSM subtyping did not find any potentially risky groups. Subtype I exhibits a greater concentration of hyperactivity and impulsivity-related symptoms and less stability in inhibition control, which may explain this association.

## 5. DISCUSSION

Multimodal clustering analysis is an essential tool for identifying meaningful subgroups beyond psychiatric diagnoses, bridging psychiatric symptoms to biological targets for a better understanding of disease progression and shedding light on future precision treatments. The ABCD study offers a comprehensive dataset for investigating multimodal integration techniques to understand the heterogeneity of adolescent development. We introduce MINDS, a Bayesian hierarchical joint model with latent variables, to integrate clinical symptoms and neuro-cognitive measures, utilizing Pólya-Gamma augmentation for posterior approximation, which facilitates Gibbs sampling. MINDS is more advanced than the alternative methods, e.g., iClusterBayes, and other two-step methods, in that it clusters on the original data instead of on intermediate features, which may lead to the loss of shared information, reducing the clustering efficiency. Extensive simulations are performed to assess the consistency of the estimators and the clustering efficiency of our method. We applied MINDS to the ABCD study, where our method demonstrates greater robustness compared to conventional clustering approaches, providing a more reliable classification of clinical subtypes. By identifying meaningful distinctions among ADHD and OCD subgroups, our findings contribute to the development of future psychological and educational interventions tailored to specific clinical presentations in mental health.

There are several challenges to MINDS. The estimation of some elements of the loading matrix can have large variances when the initial values are non-informative and the dimensions of binary items and continuous outcomes are high. Using informative initial values, for example, by fitting single-modality models, may reduce variability. As the number of items within each modality increases, computational demands increase, posing challenges in handling high-dimensional data. This can significantly slow down clustering and inference, particularly when there are many variables across modalities. Exploring variational inference methods may mitigate these computational challenges in a Bayesian approach.

Lastly, several extensions may be of interest. Introducing a penalty to the loading matrix could help manage high-dimensional data more effectively. Additionally, our method could be adapted to integrate other data types, such as zero-inflated or complex-omics data, making it even more applicable to diverse datasets in precision medicine. Validating our identified subtypes in an independent study, such as All*of*Us (All of Us Research Program Investigators, 2019) or other clinical samples, would be important.

## Data Availability

Data used in the preparation of this manuscript were obtained from the Adolescent Brain Cognitive Development (ABCD) Study (DOI: 10.15154/z563-zd24), held in the National Institute of Mental Health (NIMH) Data Archive (NDA). NDA is a collaborative informatics system created by the National Institutes of Health to provide a national resource to support and accelerate research in mental health.

## Appendix. Derivation of the Gibbs sampling steps for the joint model

The conditional probability of *y* given all the rest parameters in the joint model in Section 2.3 is

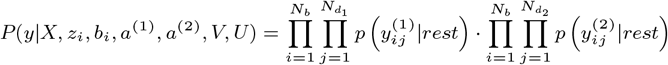

Denote the first part of the likelihood as *PartA* and the second part of the likelihood as *PartB*.

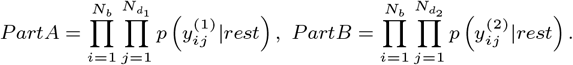

Below we will show how to derive the posterior distribution of the *t*th latent construct *X*_*t*_,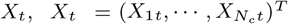. The conditional distribution of *X*_*t*_ is *P* (*X*_*t*_|*rest*) = *P* (*X*_*t*_) · *A* · *B*. Given the prior 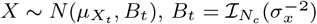, we have 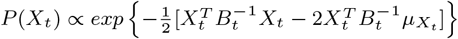.

As mentioned in section 2.3, we introduce a latent variable *ω* following the Pólya-Gamma distribution such that 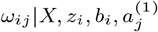, *V*_.*j*_ ∼ *PG*(1, *ψ*_*ij*_), where 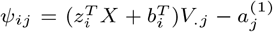. The conditional probability of *y*^(1)^ given all the rest parameters *A* can be expressed as

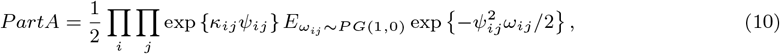

where *κ*_*ij*_ = *y*_*ij*_ − 1*/*2. Given *ω*_*ij*_, the expectation part in Equation (10) turns to be exp 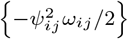, implying that the conditional probability of 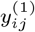 belongs to the Gaussian family.

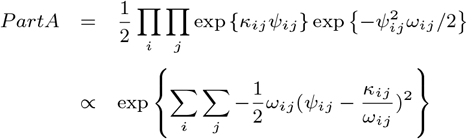

Plugging *ψ*_*ij*_, we have *PartA* ∝ exp 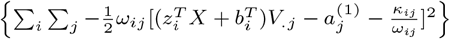. Splitting *X* into *X*_*t*_ and 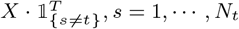, we have

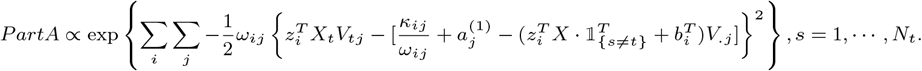

Denote 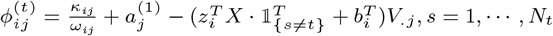,we have

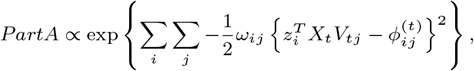

which can be written in a matrix form such as

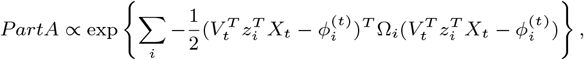

where 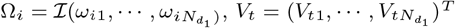 and 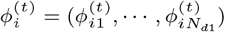.

The second part of the likelihood, *PartB*, can be expressed in a matrix form in a similar way, such that

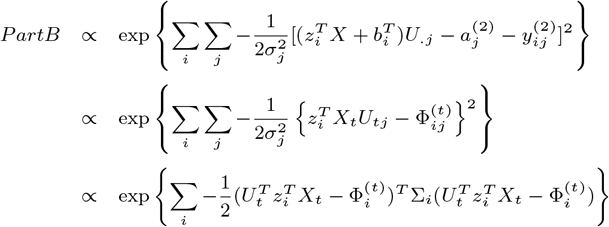

where 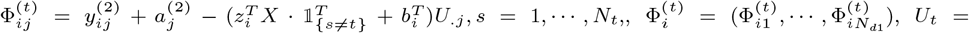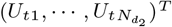, and 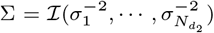. Integrating all the three matrix forms of *P* (*X*_*t*_), *PartA* and *PartB*, we have

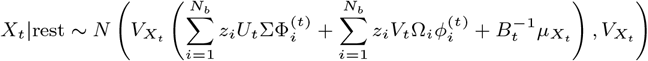

where 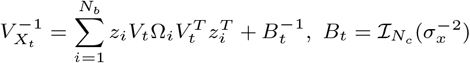.

Posterior distributions of other parameters can be derived in an analogous way.

## Competing interests

No competing interest is declared.

## Author contributions statement

Y.Z. conceived and developed the methodological framework, implemented the MINDS algorithm, performed data analysis, and drafted the manuscript. Y.W. contributed to the theoretical development, supervised the statistical modeling, and provided critical revisions. Y.L. supervised the overall study design, provided guidance on the clinical interpretation of the findings, and contributed to manuscript editing and revision. All authors reviewed and approved the final manuscript.

## Acknowledgments

Data used in the preparation of this manuscript were obtained from the Adolescent Brain Cognitive Development (ABCD) Study (DOI: 10.15154/z563-zd24), held in the National Institute of Mental Health (NIMH) Data Archive (NDA). NDA is a collaborative informatics system created by the National Institutes of Health to provide a national resource to support and accelerate research in mental health. This manuscript reflects the views of the authors and may not reflect the opinions or views of the NIH or of the Submitters submitting original data to NDA.

This work was supported by the National Institute of Mental Health [MH123487], National Institute of Neurological Disorders and Stroke [NS073671], and National Institute of Health [TL1TR001875]. The authors declare that they have no known competing financial interests or personal relationships that could have appeared to influence the work reported in this paper. This publication was neither originated nor managed by AbbVie, and it does not communicate the results of AbbVie-sponsored Scientific Research. Thus, it is not in the scope of the AbbVie Publication Procedure (PUB-100).

## Notes

### Competing Interest Statement

The authors have declared no competing interest.

### Funding Statement

https://abcdstudy.org/

### Author Declarations

The study used only openly available human data that were originally located at https://abcdstudy.org/

### Summary of Updates

compare with more alternatives, iClusterBayes and two-step JIVE ; include subjects that have missing variables in the ABCD data analysis

