## Supplementary Materials for "An Integrative Approach for Subtyping Mental Disorders Using Multimodal Data"

### 1 Supplementary figures and tables for the simulation section

Table 1: Training set performance of MINDS and alternative methods (mean(sd) across 200 replicates). Each model integrates one binary and one continuous modality (10–30 binary items; 10 continuous measures).

| Training Size | Items | MINDS | iCluster | Two-step JIVE | K-means | Hclust | Two-step K-means | Two-step Hclust |
|---|---|---|---|---|---|---|---|---|
| **Training Set Classification Error** | | | | | | | | |
| 1000 | 10 | 0.119(0.072) | 0.260(0.023) | 0.347(0.026) | 0.268(0.102) | 0.371(0.028) | 0.260(0.100) | 0.322(0.074) |
| 2000 | 10 | 0.112(0.059) | 0.251(0.018) | 0.358(0.014) | 0.304(0.084) | 0.367(0.026) | 0.285(0.091) | 0.348(0.072) |
| 1000 | 20 | 0.075(0.059) | 0.217(0.036) | 0.450(0.053) | 0.211(0.155) | 0.379(0.069) | 0.238(0.098) | 0.291(0.070) |
| 2000 | 20 | 0.077(0.059) | 0.197(0.031) | 0.450(0.067) | 0.211(0.156) | 0.376(0.059) | 0.255(0.091) | 0.267(0.071) |
| 1000 | 30 | 0.095(0.038) | 0.245(0.040) | 0.475(0.016) | 0.232(0.139) | 0.311(0.029) | 0.267(0.091) | 0.287(0.064) |
| 2000 | 30 | 0.102(0.046) | 0.247(0.039) | 0.492(0.012) | 0.260(0.133) | 0.311(0.030) | 0.256(0.091) | 0.338(0.085) |
| **Training Set Jaccard Distance** | | | | | | | | |
| 1000 | 10 | 0.263(0.074) | 0.511(0.026) | 0.620(0.014) | 0.456(0.072) | 0.518(0.016) | 0.448(0.076) | 0.538(0.070) |
| 2000 | 10 | 0.258(0.065) | 0.503(0.017) | 0.638(0.009) | 0.483(0.063) | 0.512(0.014) | 0.459(0.062) | 0.554(0.066) |
| 1000 | 20 | 0.194(0.072) | 0.383(0.032) | 0.633(0.047) | 0.351(0.142) | 0.556(0.058) | 0.396(0.076) | 0.488(0.078) |
| 2000 | 20 | 0.191(0.071) | 0.350(0.026) | 0.621(0.056) | 0.348(0.144) | 0.555(0.048) | 0.385(0.072) | 0.486(0.111) |
| 1000 | 30 | 0.207(0.041) | 0.414(0.032) | 0.723(0.008) | 0.351(0.123) | 0.415(0.022) | 0.411(0.079) | 0.434(0.084) |
| 2000 | 30 | 0.214(0.049) | 0.410(0.031) | 0.735(0.004) | 0.382(0.124) | 0.421(0.032) | 0.400(0.080) | 0.508(0.111) |

### 2 Supplementary figures and tables for real data analysis

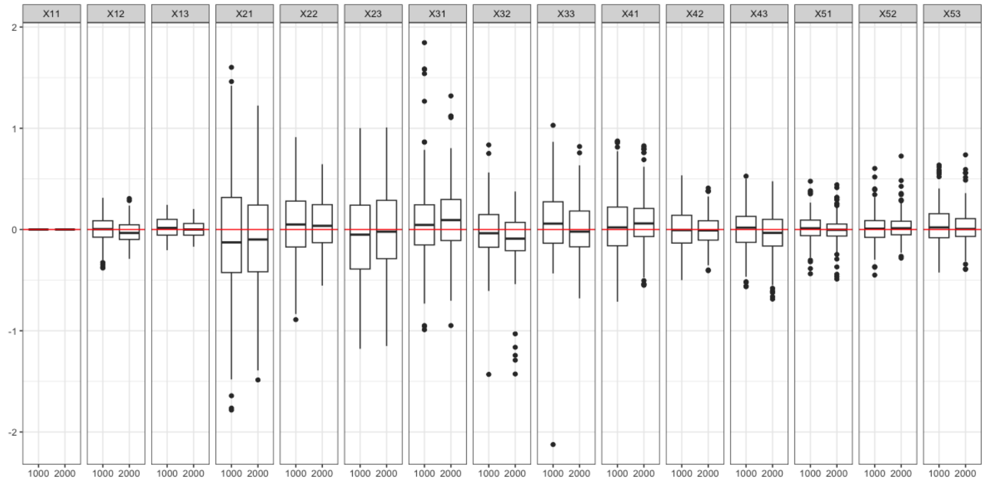


Figure 1: Estimation of cluster center $X$; results are from a joint model for integrating one binary modality with 10 binary items and one continuous modality with 10 continuous measures based on 200 replicates; simulations are conducted for 1,000 vs. 2,000 sample size; red lines represent true values.

Table 2: Mean(sd) of the testing set classification error and Jaccard distance of MINDS and alternative methods based on 200 replicates. Training sample sizes ranged from 1,000 to 2,000 subjects, with models integrating one binary and one continuous modality (10–30 binary items and 10 continuous measures).

| **Items** | **Training size** | **MINDS** | **Two-step JIVE** | **K-means** | **Hclust** | **Two-step K-means** | **Two-step Hclust** |
|---|---|---|---|---|---|---|---|
| | | | **Testing Set Classification Error** | | | | |
| 10 | 1000 | 0.178(0.103) | 0.398(0.073) | 0.303(0.096) | 0.359(0.037) | 0.260(0.112) | 0.335(0.098) |
| 10 | 2000 | 0.151(0.080) | 0.394(0.072) | 0.286(0.103) | 0.355(0.036) | 0.253(0.114) | 0.311(0.088) |
| 20 | 1000 | 0.108(0.119) | 0.392(0.167) | 0.249(0.168) | 0.319(0.054) | 0.211(0.125) | 0.253(0.075) |
| 20 | 2000 | 0.102(0.110) | 0.362(0.167) | 0.245(0.176) | 0.313(0.055) | 0.209(0.132) | 0.228(0.064) |
| 30 | 1000 | 0.240(0.087) | 0.398(0.094) | 0.256(0.136) | 0.342(0.038) | 0.241(0.107) | 0.312(0.055) |
| 30 | 2000 | 0.210(0.054) | 0.404(0.091) | 0.249(0.138) | 0.338(0.034) | 0.236(0.111) | 0.296(0.069) |
| | | | **Testing Set Jaccard Distance** | | | | |
| 10 | 1000 | 0.398(0.051) | 0.632(0.033) | 0.481(0.068) | 0.504(0.010) | 0.441(0.076) | 0.540(0.054) |
| 10 | 2000 | 0.380(0.042) | 0.632(0.030) | 0.467(0.073) | 0.504(0.011) | 0.435(0.079) | 0.532(0.054) |
| 20 | 1000 | 0.195(0.083) | 0.548(0.091) | 0.375(0.157) | 0.476(0.049) | 0.333(0.107) | 0.419(0.057) |
| 20 | 2000 | 0.189(0.080) | 0.530(0.098) | 0.368(0.163) | 0.465(0.047) | 0.329(0.113) | 0.401(0.050) |
| 30 | 1000 | 0.481(0.052) | 0.549(0.057) | 0.376(0.123) | 0.449(0.038) | 0.371(0.099) | 0.437(0.037) |
| 30 | 2000 | 0.477(0.037) | 0.557(0.051) | 0.363(0.121) | 0.445(0.033) | 0.373(0.101) | 0.434(0.042) |

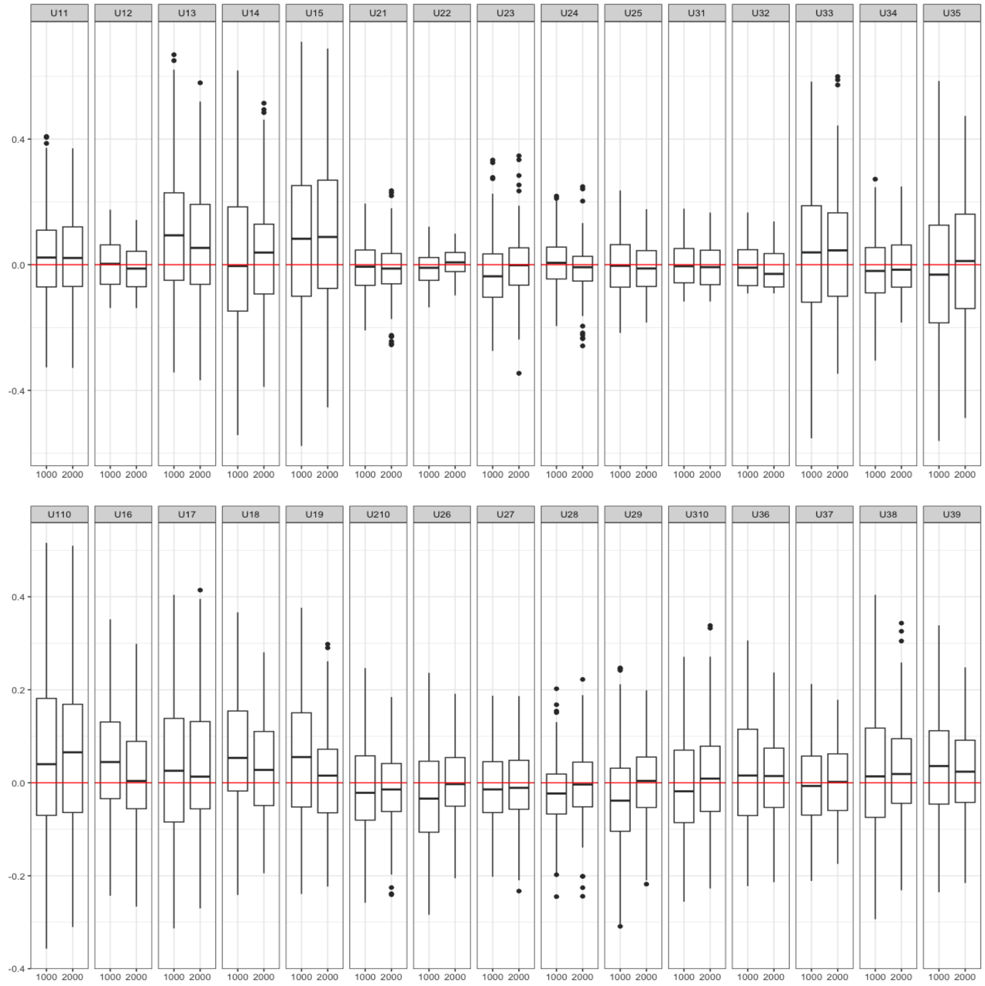


Figure 2: Estimation of Loading matrix $U$ for continuous measures; results are from a joint model for integrating one binary modality with 10 binary items and one continuous modality with 10 continuous measures based on 200 replicates; simulations are conducted for 1,000 vs. 2,000 sample size; red lines represent true values.

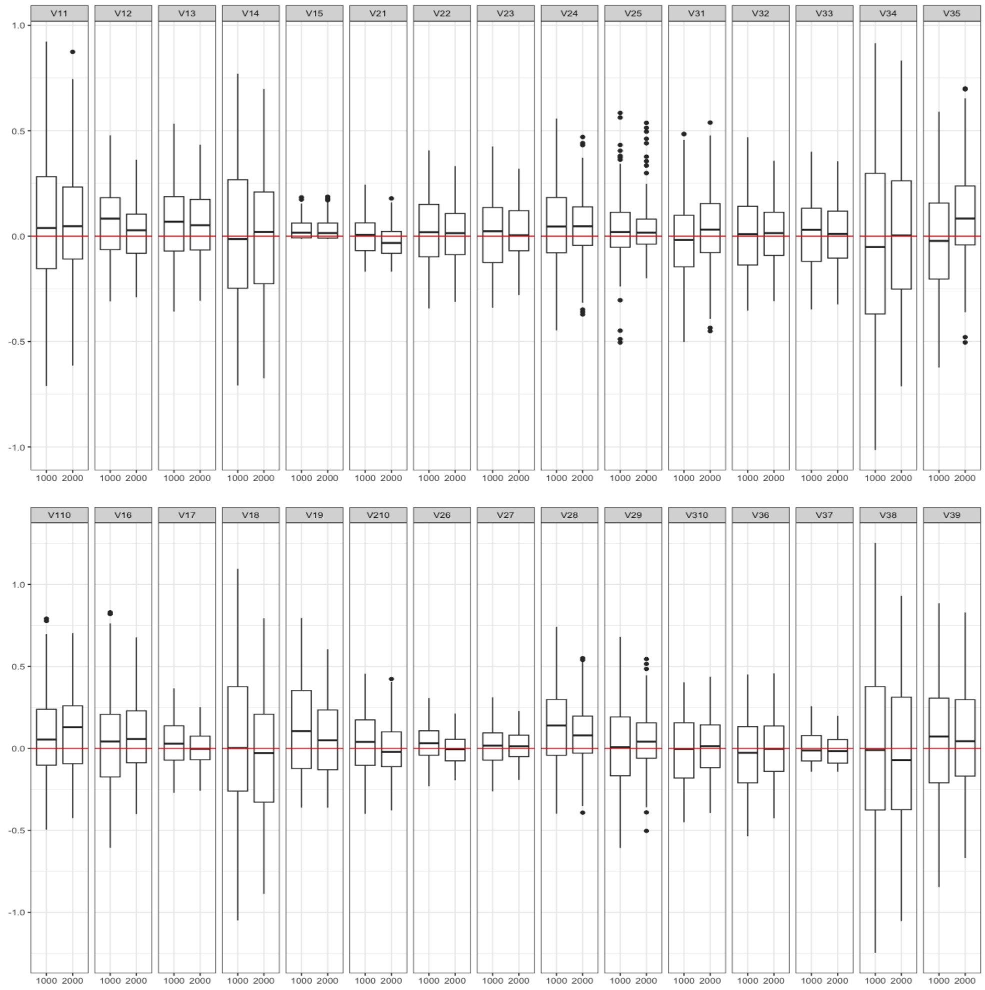


Figure 3: Estimation of Loading matrix $V$ for binary items; results are from a joint model for integrating one binary modality with 10 binary items and one continuous modality with 10 continuous measures based on 200 replicates; simulations are conducted for 1,000 vs. 2,000 sample size; red lines represent true values.

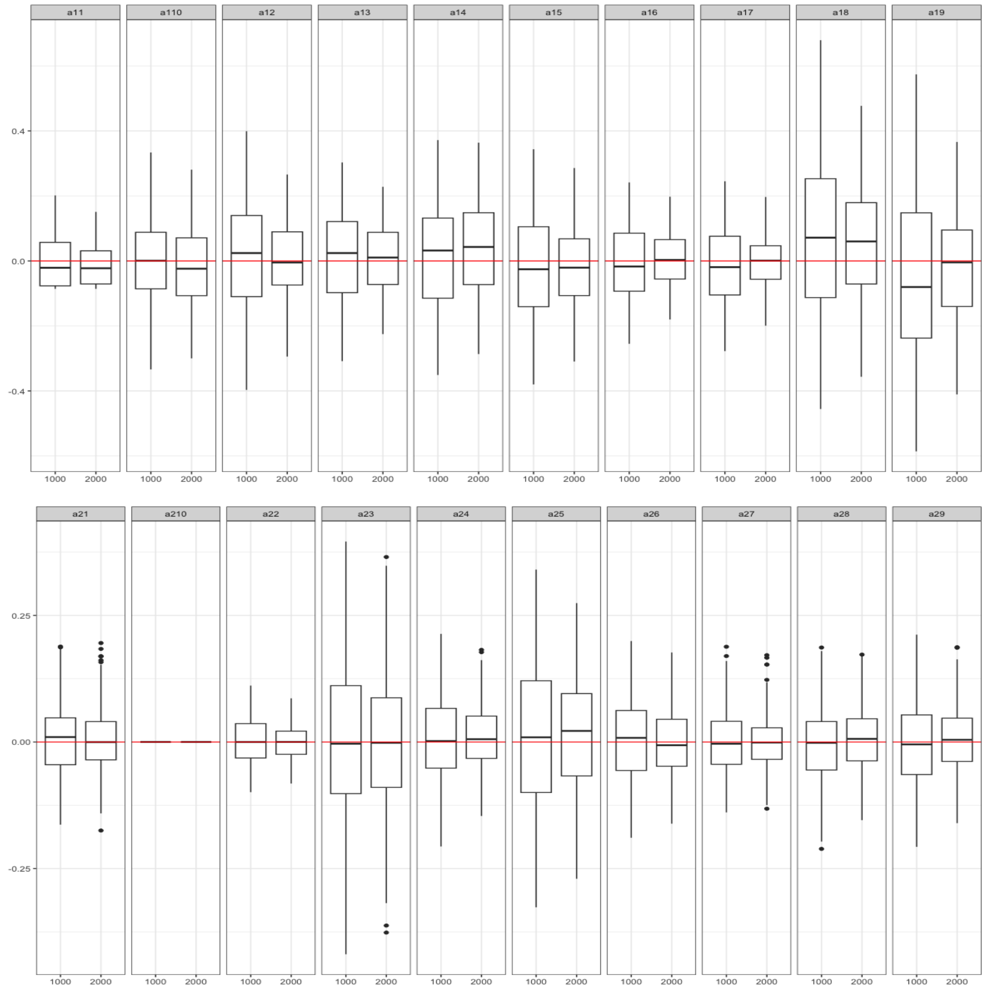


Figure 4: Estimation of item difficulty $a_1$ for binary items and $a_2$ for continuous measures; results are from a joint model for integrating one binary modality with 10 binary items and one continuous modality with 10 continuous measures based on 200 replicates; simulations are conducted for 1,000 vs. 2,000 sample size; red lines represent true values.

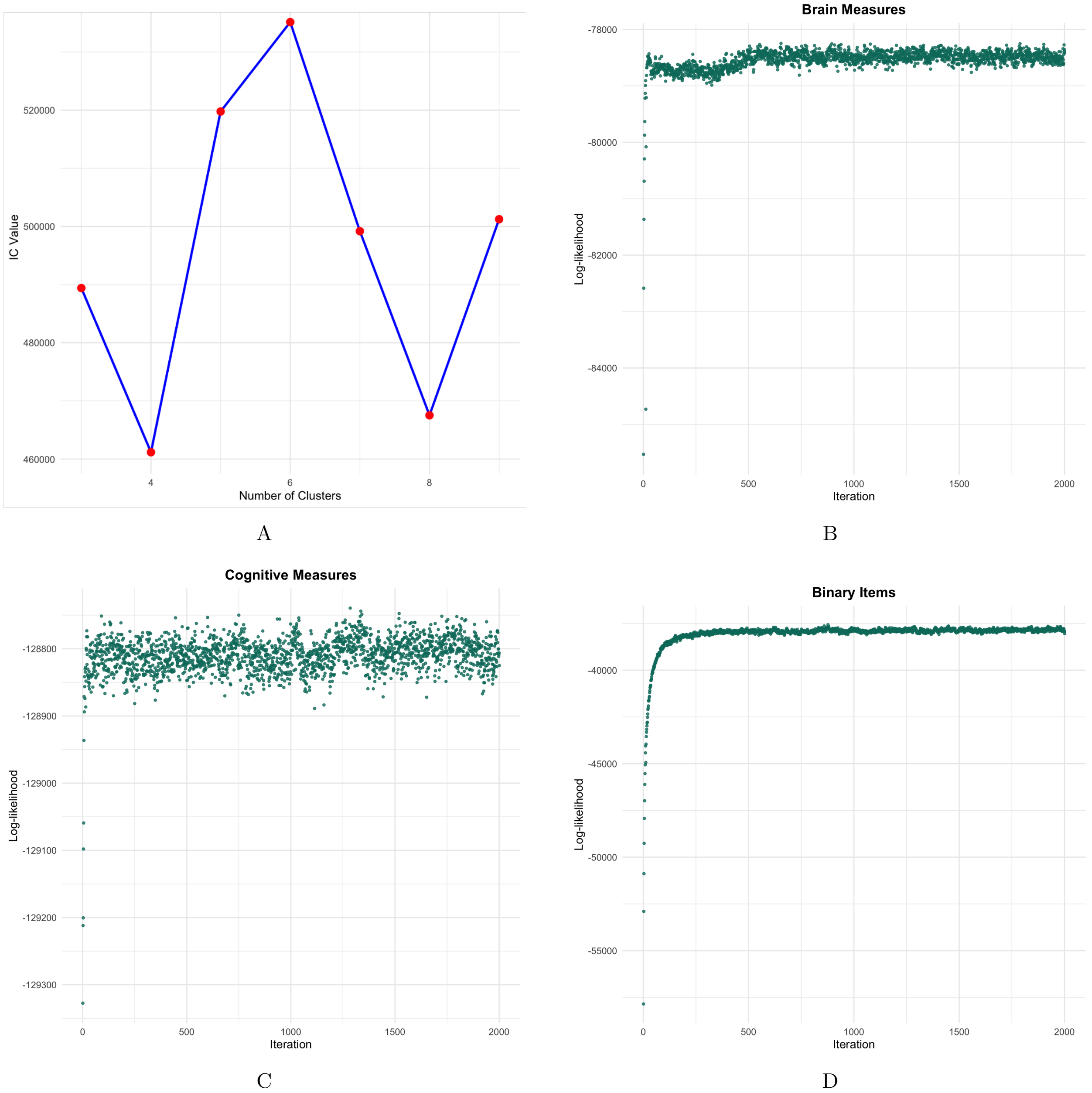


Figure 5: **Model selection and log-likelihood trace plot in each modality for the real data analysis.** (A) IC value at $k = 3, 4, 5, 6, 7, 8, 9$. The model fits best when $k = 4$. (B-D) Posterior log-likelihood of each modality when $k = 4$.
